# Pediatric brain tumor classification using deep learning on MR-images with age fusion

**DOI:** 10.1101/2024.09.05.24313109

**Authors:** Iulian Emil Tampu, Tamara Bianchessi, Ida Blystad, Peter Lundberg, Per Nyman, Anders Eklund, Neda Haj-Hosseini

## Abstract

**Purpose:** To implement and evaluate deep learning-based methods for the classification of pediatric brain tumors in MR data.

**Materials and methods:** A subset of the “Children’s Brain Tumor Network” dataset was retrospectively used (n=178 subjects, female=72, male=102, NA=4, age-range [0.01, 36.49] years) with tumor types being low-grade astrocytoma (n=84), ependymoma (n=32), and medulloblastoma (n=62). T1w post-contrast (n=94 subjects), T2w (n=160 subjects), and ADC (n=66 subjects) MR sequences were used separately. Two deep-learning models were trained on transversal slices showing tumor. Joint fusion was implemented to combine image and age data, and two pre-training paradigms were utilized. Model explainability was investigated using gradient-weighted class activation mapping (Grad-CAM), and the learned feature space was visualized using principal component analysis (PCA).

**Results:** The highest tumor-type classification performance was achieved when using a vision transformer model pre-trained on ImageNet and fine-tuned on ADC images with age fusion (MCC: 0.77 ± 0.14 Accuracy: 0.87 ± 0.08), followed by models trained on T2w (MCC: 0.58 ± 0.11, Accuracy: 0.73 ± 0.08) and T1w post-contrast (MCC: 0.41 ± 0.11, Accuracy: 0.62 ± 0.08) data. Age fusion marginally improved the model’s performance. Both model architectures performed similarly across the experiments, with no differences between the pre-training strategies. Grad-CAMs showed that the models’ attention focused on the brain region. PCA of the feature space showed greater separation of the tumor-type clusters when using contrastive pre-training.

**Conclusion:** Classification of pediatric brain tumors on MR-images could be accomplished using deep learning, with the top-performing model being trained on ADC data, which is used by radiologists for the clinical classification of these tumors.

**Key points:**

- The vision transformer model pre-trained on ImageNet and fine-tuned on ADC data with age fusion achieved the highest performance, which was significantly better than models trained on T2w (second-best) and T1w-Gd data.
- Fusion of age information with the image data marginally improved classification, and model architecture (ResNet50 -vs -ViT) and pre-training strategies (supervised -vs -self-supervised) did not show to significantly impact models’ performance.
- Model explainability, by means of class activation mapping and principal component analysis of the learned feature space, show that the models use the tumor region information for classification and that the tumor type clusters are better separated when using age information.

**Summary:** Deep learning-based classification of pediatric brain tumors can be achieved using single-sequence pre-operative MR data, showing the potential of automated decision support tools that can aid radiologists in the primary diagnosis of these tumors.

## Introduction

Tumors in the central nervous system are the second most common type of cancer in children and young adults up to the age of 19, with an estimated age-standardized rate (in 100,000 population) of 1.2 for incidence and 0.60 for mortality worldwide^1^, where brain tumors account for about 57% of the total causes of cancer deaths in this population^2^. Pediatric brain tumors (PBT) can be grouped concerning the location relative to the tentorium, as infratentorial or supratentorial. Tumors in the infratentorial brain region (posterior fossa) are more common in pediatric patients; however, the frequency varies depending on age^3–5^. Brain tumor treatment procedures are usually complicated where tumor detection and preliminary diagnosis are based on magnetic resonance images (MRI), and treatment planning also uses histopathological and molecular analysis of the tissue sample^6^. Diagnosis by radiologists, when comparing the first MRI diagnosis to the final histology diagnosis, varies greatly among tumor types and locations, with an overall sensitivity of 72% for broad tumor type classification (range 0-100%), which shows the need for computational methods to improve qualitative assessments^7^. Deep learning algorithms have been successfully applied to several medical image-related tasks and can be trained to assist radiologists in diagnosing brain tumors based on MR. Even though deep learning methods have led to reasonable advancements in adult brain tumor detection, classification, and segmentation^8–10^, their implementation in pediatric cases has been limited^11,12^ mainly due to the lack of large and standardized open access datasets^13,14^. Deep learning models trained on MR-images from adults will not perform well on images from children, since PBTs have different diagnostic properties. The “Children’s Brain Tumor Network” (CBTN)^15,16^ is one of the largest PBT datasets, and could potentially be used in the future similarly to the adult brain tumor segmentation challenge (BraTS)^17–19^, as a standard and reference dataset for development and comparison of deep learning methods. This study is, to the best of our knowledge, the first report on the implementation of deep learning on the MR dataset from CBTN for brain tumor classification, and also one of few hitherto published MR-based deep learning studies on any brain tumor pediatric dataset.

This exploratory study aimed to investigate deep learning-based methods for the classification of pediatric brain tumors, considering different pre-operative MRI sequences, and fusing age and image information. A convolutional neural network (ResNet50) and a vision transformer (ViT) were implemented and evaluated, exploring three pre-training strategies, and investigating model explainability by visualization of activation maps and the learned feature space.

## Material and Methods

### Dataset cohort

In this retrospective study, the dataset was obtained upon application to and approval from CBTN^15^ (accessed in 2021). The downloaded dataset contained 326 subjects, with tumor type information available for 273 subjects (females=153, males=116, not-available=4, age-range [0.01, 36.49] years). Patients older than 18 years (n=3) were included in the dataset given the pediatric tumor type diagnosis. The tumor types available were low-grade astrocytoma (ASTR) (n=132), medulloblastoma (MB) (n=67), ependymoma (EP) (n=45), atypical teratoid rhabdoid tumor (ATRT) (n=20), diffuse intrinsic pontine glioma (DIPG) (n=6), ganglioglioma (n=1), germinoma (n=1), and teratoma (n=1). Due to the limited number of subjects for DIPG, ganglioglioma, germinoma, and teratoma categories, these tumor types were excluded from the subsequent analysis. From the remaining tumor type groups, T1w-Gd, T2w MR sequences and diffusion-weighted (DW-MR) data were collected and used in the analysis.

### Data selection and exclusion

An automated selection based on the image quality followed by a visual assessment was performed. Quality selection for T1w-Gd and T2w data was based on the voxel resolution, removing those with axial in-plane resolution larger then 1mm, and with less than 50 axial slices. These values were chosen to avoid artifacts due to a low image resolution. For the diffusion-weighted data, scans with at less than six diffusion-encoding directions were excluded. Visual assessment of all individual images was performed and data were excluded if: (i) images were acquired post-operatively, (ii) images showed the spine only, (iii) the tumor was not visible, (iv) the transversal plane had been clipped, and (v) image artifacts (motion, metal, induced by neurosurgical clips) were present. By visual assessment, the tumor location was saved as a boundary box.

### Pre-processing of image and age data

The DW-MR data was processed using MRtrix3 software^20^ to obtain diffusion tensors from which the ADC map was calculated. Brain extraction was performed, followed by data harmonization, using a per-sequence voxel intensity normalization and interpolation down to 1 mm isotropic resolution. These steps were performed since the CBTN dataset was collected on a variety of MR scanners (manufacturer, field strength, gradient performance, etc.)^16^. The final volumes were reshaped to have 224 × 224 pixels in the transverse plane. Transversal 2D slices positioned within 20-80% of the tumor boundary box were extracted from the volumetric data to ensure that images showing only small portions of the tumor were not included. Transversal slices were used instead of the volumetric data due to the limited number of subjects available for training 3D deep learning models. A detailed description of the pre-processing steps and software used is available in the *Supplementary material*. The age in days of each subject from the earliest available scan was obtained from the CBTN portal, converted in years, and normalized using *z-score* normalization using the [0.5^th^, 99.5^th^] value range. The final composition of the dataset with age and sex information is summarized in Table 1.

**Table 1.**
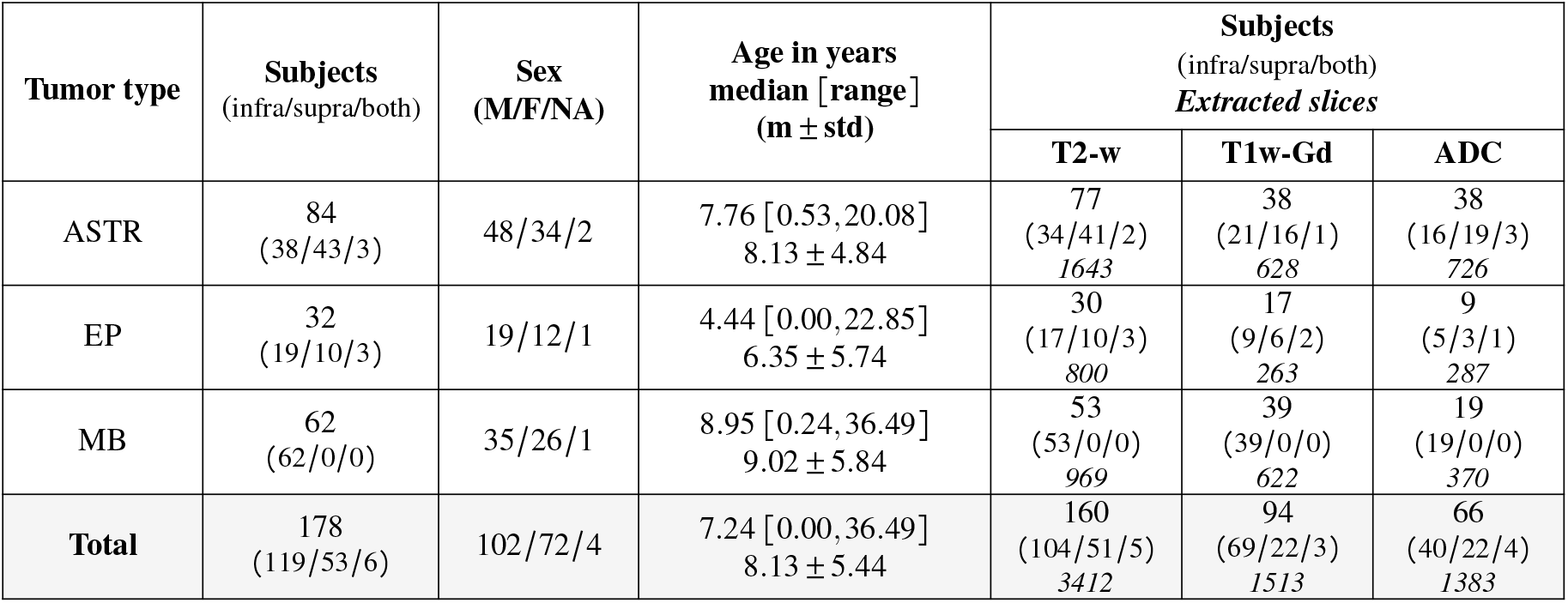
Per tumor type and MR-sequence summary of the dataset. The age information was obtained from the earliest scan available for each subject. The number of extracted slices reflects the slices within the 20-80% of the tumor boundary box. m: mean, std: standard deviation, M/F: male/female, NA: not available, ASTR: astrocytoma, EP: ependymoma, MED: medulloblastoma.

| Tumor type | Subjects<br>(infra/supra/both) | Sex<br>(M/F/NA) | Age in years<br>median [range]<br>(m ± std) | Subjects<br>(infra/supra/both)<br>*Extracted slices* | | |
| --- | --- | --- | --- | --- | --- | --- |
|  |  |  |  | T2-w | T1w-Gd | ADC |
| ASTR | 84<br>(38/43/3) | 48/34/2 | 7.76 [0.53, 20.08]<br>8.13 ± 4.84 | 77<br>(34/41/2)<br>1643 | 38<br>(21/16/1)<br>628 | 38<br>(16/19/3)<br>726 |
| EP | 32<br>(19/10/3) | 19/12/1 | 4.44 [0.00, 22.85]<br>6.35 ± 5.74 | 30<br>(17/10/3)<br>800 | 17<br>(9/6/2)<br>263 | 9<br>(5/3/1)<br>287 |
| MB | 62<br>(62/0/0) | 35/26/1 | 8.95 [0.24, 36.49]<br>9.02 ± 5.84 | 53<br>(53/0/0)<br>969 | 39<br>(39/0/0)<br>622 | 19<br>(19/0/0)<br>370 |
| **Total** | 178<br>(119/53/6) | 102/72/4 | 7.24 [0.00, 36.49]<br>8.13 ± 5.44 | 160<br>(104/51/5)<br>3412 | 94<br>(69/22/3)<br>1513 | 66<br>(40/22/4)<br>1383 |

### Network architecture and training

Two deep learning model architectures extensively used in literature were employed in this study, distinguished by their feature extraction approach: ResNet50^21^ and Vision Transformer^22^ (ViT), in its *base 16* version. ResNet50^21^ is a deep convolutional neural network that uses stacked 2D convolutional layers and residual connections to extract image features. ViT^22^ is a transformer-based model free from convolution operations that uses self-attention to learn local and global relations between non-overlapping patches in an image. Both methods serve as image feature extractors, producing a 1D representation of an input image suitable for classification, with ViT showing to perform better than ResNet-like models on natural image classification tasks as well as being more robust to image perturbations when trained on sufficient data^23^. Given the limited training data available, transfer learning was used with the image encoding models fine-tuned on the target CBTN dataset starting from pre-trained weights. Three distinct pre-training strategies were investigated: supervised pre-training on *out-of-domain* data (ImageNet1K^24^), self-supervised pre-training on *close-to-domain* data (BraTS^17–19^) and self-supervised pre-training on *in-domain* (CBTN) data. For the self-supervised pre-training, the SimCLR^25^ framework was employed (see *Pre-training* section in the *Supplementary materials* for details). We also investigated the integration of image and age through a joint fusion approach. In this case, ResNet50 and ViT models were used to encode the image data, while a tabular network encoded the age information. Figure 1 shows a schematic representation of the network architecture when trained on image and age information. For the details on the implementation, model pre-training and fine-tuning, and data augmentation, see the *Supplementary materials*.

**Figure 1.**
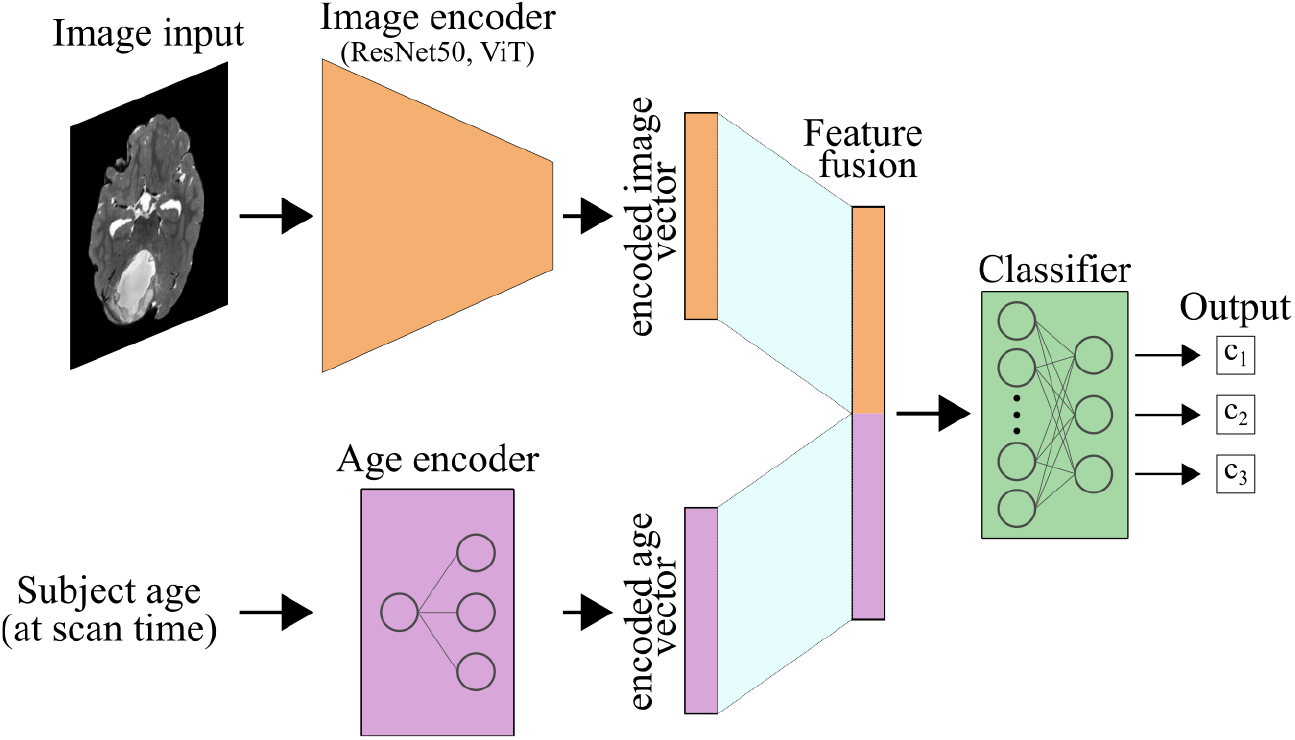
Schematic representation of image and age encoders whose 1D representations are concatenated for the final classification. ViT: visual transformer.

### Evaluation metrics and statistical methods

A ten-times repeated five-fold stratified cross-validation scheme was employed in all experiments to account for the small size of the dataset. For each of the repetitions, subject-wise splitting was performed to obtain training, validation, and testing sets. Models’ performance was evaluated volume-wise in terms of Matthew’s correlation coefficient [-1, 1]^26^ since it is a more stable metric in case of class imbalance. Accuracy and area under the ROC curve (AUC) were also computed to allow comparison with previous studies. Class-wise F1-score, precision, and recall were additionally computed. Volume-wise predictions were obtained by soft voting aggregation of the models’ predicted probability for each of the slices in a volume. The Wilcoxon signed-rank test (two-sided) was used to investigate if there were difference in classification performance between models trained on image data alone or fused with age information, or when using different pre-training strategies. The Wilcoxon rank-sums (two-sided) test was instead used to compare models when trained on different MR sequences. A *p*-value< 0.05 was considered significant with Bonferroni correction applied when multiple comparisons were performed.

### Model explainability and learned feature space visualization

In this work, Grad-CAM^27,28^ were computed for the last convolutional layer of the ResNet50 models, and for the last attention block in the ViT models, for the ground truth class. Grad-CAMs were employed to ensure that the models focused on relevant regions of the input image for classification, rather than to elucidate the specific reasons or features used by models for prediction. Additionally, principal component analysis (PCA) was performed on the image feature vectors obtained from the trained models to visualize the effect of pre-training and image-age fusion.

## Results

### Classification performance

The highest classification performance was achieved by the ViT model pre-trained on ImageNet and fine-tuned on ADC data with age fusion (MCC: 0.77 ± 0.14, Accuracy: 0.87 ± 0.08). This was significantly higher than the best-performing models trained on either T2w (MCC: 0.58 ± 0.11) or T1w-Gd (MCC: 0.45 ± 0.16) data. Class-wise performance (see *Supplementary material* Table S1), showed that the classification of EP is the most challenging across settings, with an average F1-score over all the experiments of 0.37 ± 0.28, while ASTR and MED obtained 0.74 ± 0.18 and 0.76 ± 0.15, respectively. Looking at the overall effect of fusing image and age information, the models’ performance did not significantly change compared to models trained on image data only, except for the ResNet50 model pre-trained on ImageNet and fine-tuned on ADC data where the addition of age information significantly decreased classification performance. The effect of the three pre-training strategies was not consistent across MR sequences, model architectures, or input configuration. Moreover, there was no clear benefit between pre-training on *close-to-domain* or *in-domain* data. Of notice, the ViT models trained on ADC data had a significantly better performance when fine-tuned from ImageNet pre-trained weights compared to contrastive pre-training. Finally, looking at the different model architectures, ResNet50 and ViT models performed similarly when considering pre-training strategies and input configuration. A summary of the classification performance for all the experiments is presented in Figure 2 and in Table 2.

**Table 2.**
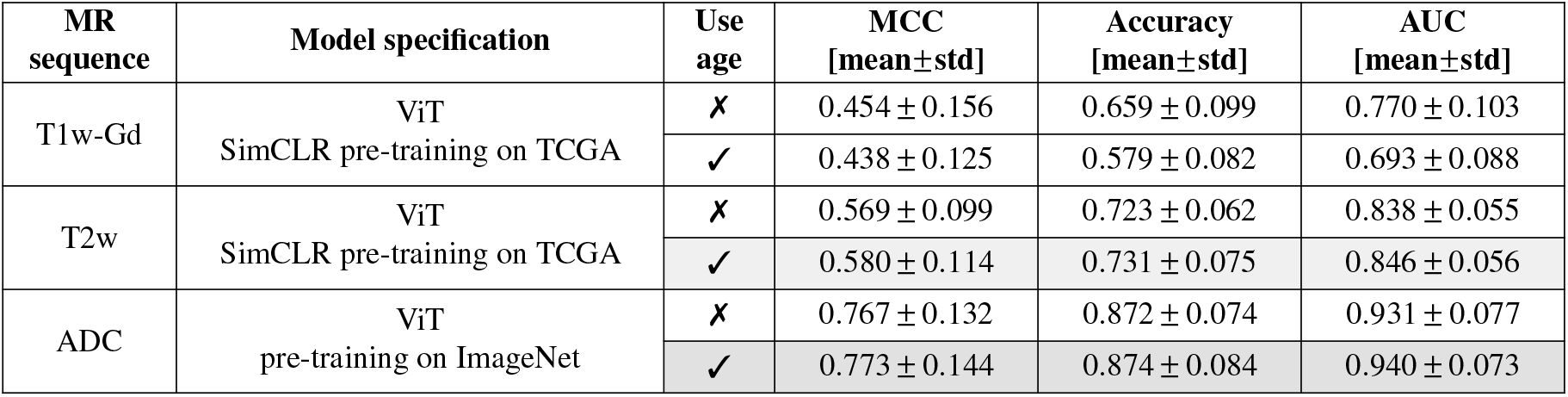
Subject-wise classification performance for the best performing models on all the available MR sequences (with and without age fusion). The overall best performing model is highlighted in gray. Models fine-tuned on ADC data perform significantly better than models fine-tuned on either T2w data or T1w-Gd. The addition of the age information did not significantly improve models’ performance. SimCLR: self-supervised contrastive pre-training strategy, std: standard deviation, MCC: Matthew’s correlation coefficient, AUC: area under the Receiver operating characteristic curve (macro-average).

**Figure 2.**
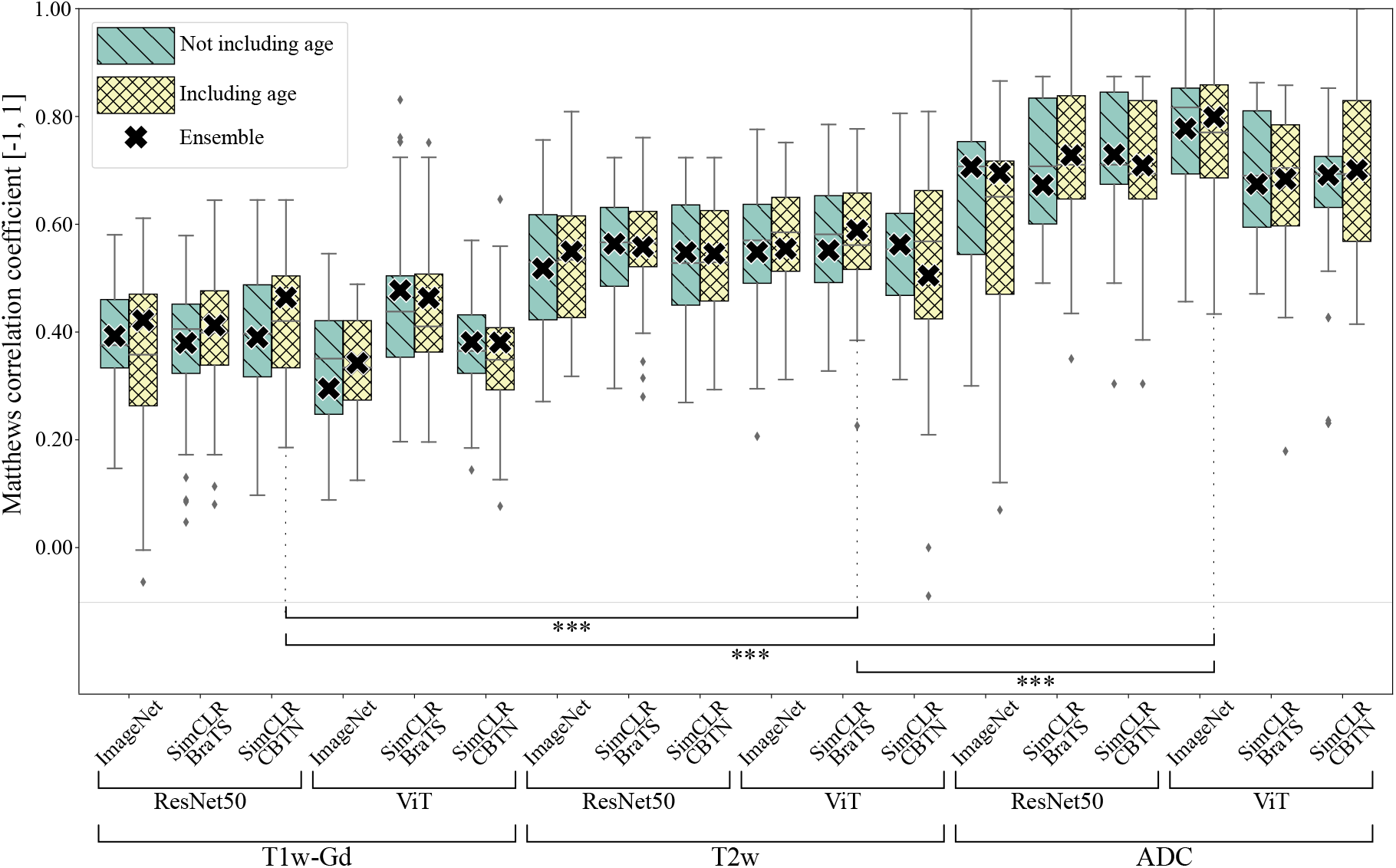
Subject-wise classification performance on the test set for all the available MR sequences (with and without age fusion) and investigated model architectures and pre-training strategies. Each box plot summarises the Matthew’s correlation coefficient for the 50 models trained thought a ten-times repeated five-fold cross-validation scheme. Outliers are shown as diamond (⧫). Statistical significance is shown for the best performing models on each MR sequence (^***^ two-sided *p*-value < 0.0001 using Wilcoxon rank-sums test adjusted with *post-hoc* Bonferroni correction). See Table 2 for the performance details of the best models for each MR sequence.

### Qualitative analysis of the image feature space

Scatter plots for features extracted using the ViT model from T2w and ADC images are shown in Figure 3. On the training set, the features of the different tumor types are grouped in distinct clusters for both T2w and ADC. The different pre-training strategies did not substantially impact the feature space, with the SimCLR pre-training on CBTN showing a marginally improved cluster separation. The fusion of the age information with the image data resulted in a larger separation between the tumor type clusters compared to when only image information was used. On the test set, clusters were less distinct, with EP features largely overlapping with ASTR and MED, reflecting the lower F1-score for this class. Scatter plots for the ResNet50 models and T1w-Gd sequence are available in the *Supplementary material* Figures S1 and S2.

**Figure 3.**
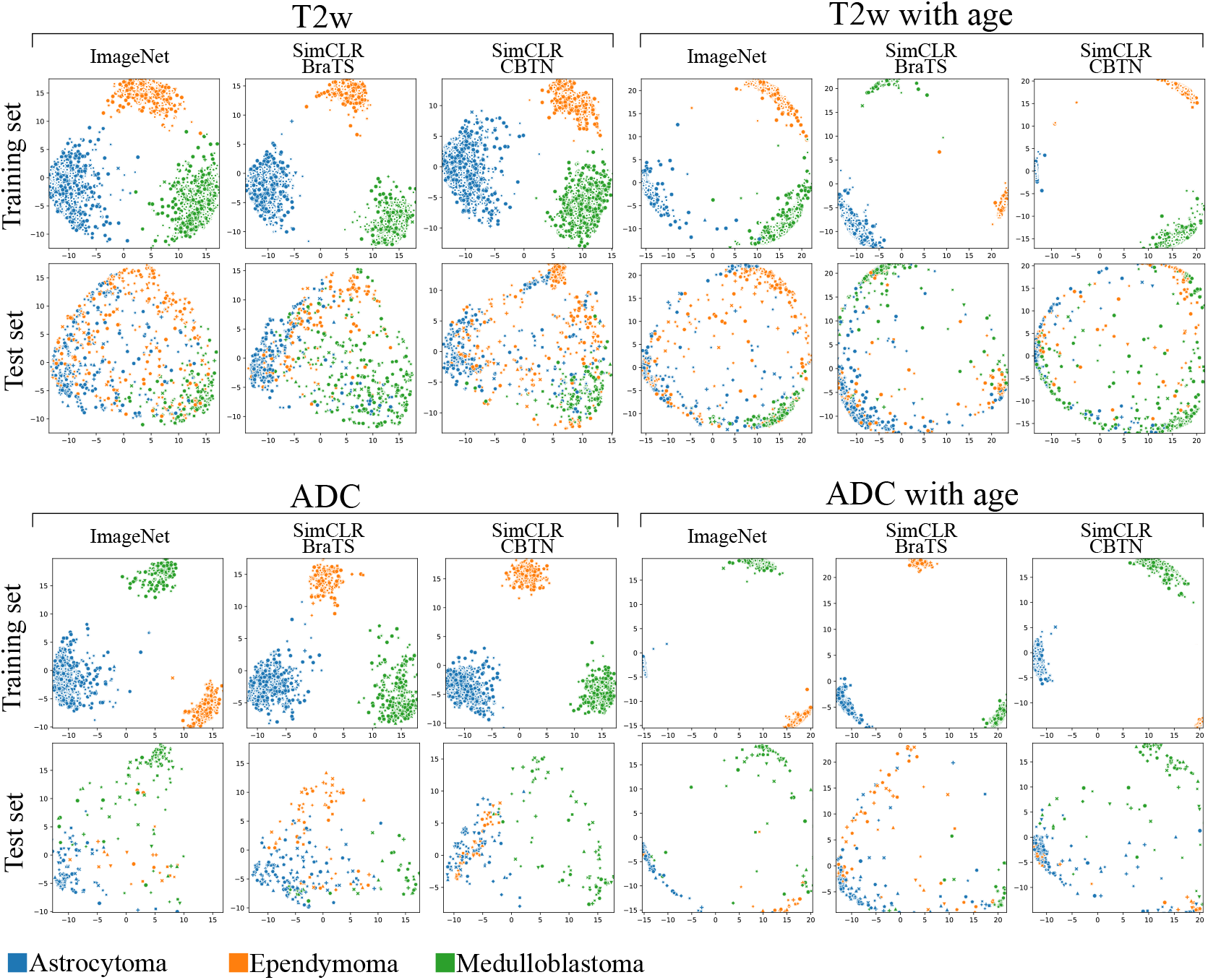
Principal component analysis (PCA) of image features extracted by ViT models fine-tuned on T2w or ADC data, with and without age information, using ImageNet or SimCLR pre-trained weights. The first and second principal components are presented, for both training and testing sets. Classes are color-coded. The addition of the age information stretches the feature space and helps, in the training set, in clustering the tumor types separately. On the test set, ependymoma samples (orange dots) are scattered and overlapping with the other two classes, confirming the low F1-score for this class. ADC: apparent diffusion coefficient, SimCLR: self-supervised contrastive pre-training.

### Grad-CAMs

Representative Grad-CAMs for models trained on the available MR sequences, with and without age fusion, and for the three pre-training strategies are presented in Figure 4. Results are shown for a transversal slice of a test subject for which all MR sequences were available. For the ResNet50 models, the Grad-CAMs focused primarily on the brain region with those of models trained on T2w data showing a better localization of the tumor compared to T1w-Gd or ADC. The Grad-CAMs of the ViT models highlighted the whole brain region with no discrimination of the tumor region, which is a consequence of the short and long relations between the image regions that these models learn. There is no overall difference in the Grad-CAMs between pre-training strategies, and when using age and image information. This was true for both the models, except for the ViT, where examples of activation being around the brain region can be found. Additional Grad-CAMs are available in the *Supplementary materials* Figure S3.

**Figure 4.**
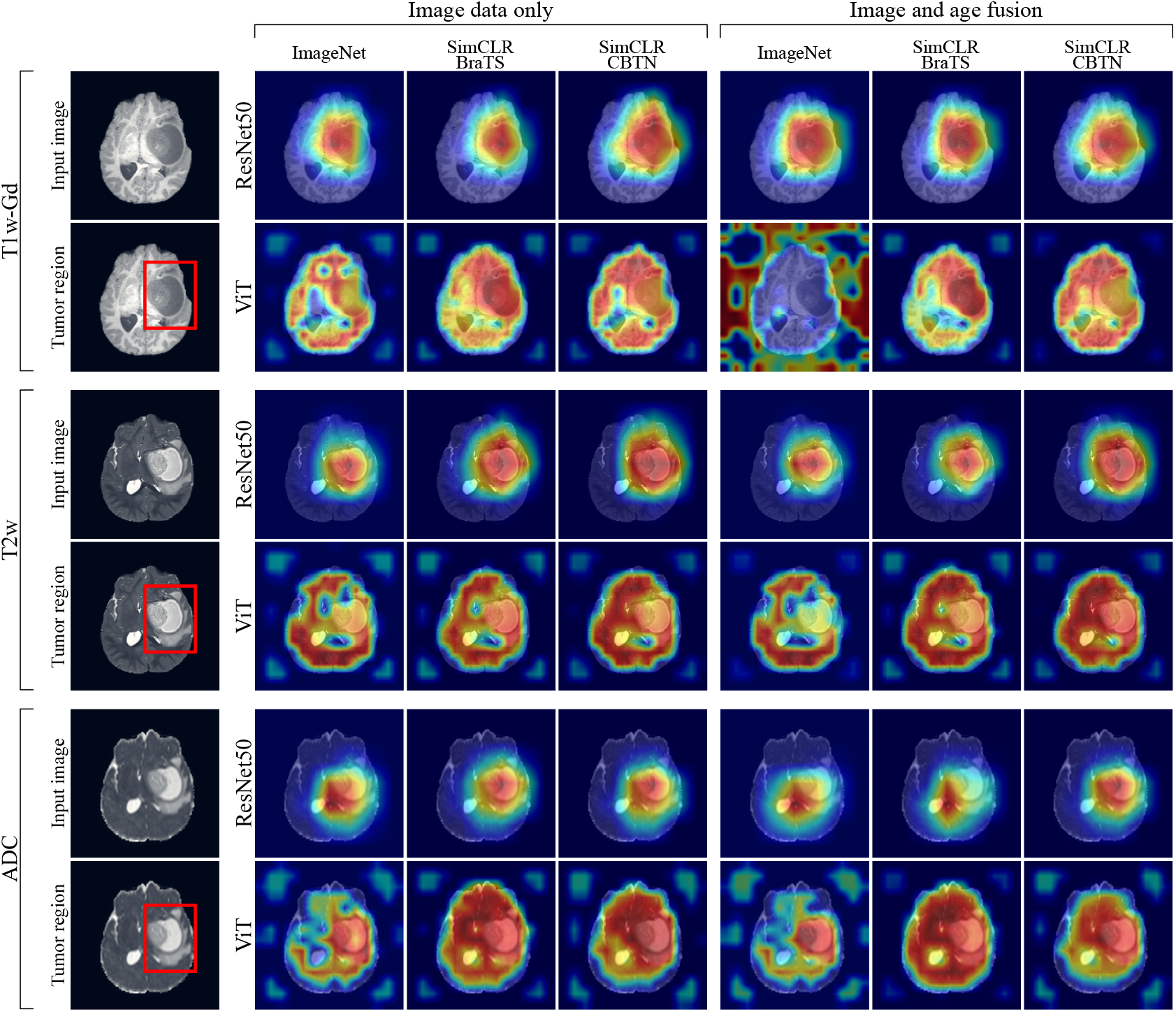
Grad-CAMs for the models trained on different MR-sequences, with and without age fusion, and for the three pre-training strategies investigated. Grad-CAMs are computed with respect to the ground truth class and for the same subject (the transversal slice was taken to be as close as possible in all MR modalities). The red square in the *tumor region* panel delineates the tumor. In the Grad-CAMs images, the red color identifies the parts of the input image used mostly contributing to the classification. ADC: apparent diffusion coefficient, SimCLR: self-supervised contrastive pre-training. ViT: visual transformer.

## Discussion

In this study, deep learning methods were implemented for the classification of PTBs based on pre-operative MR-images from the CBTN dataset. The effect of network architectures, pre-training, MR sequence, and fusion of patient age were investigated.

### Network architecture

The ResNet50 model was chosen given previous reports in literature for similar tasks on both pediatric^11,12^ and adult brain tumor datasets^29,30^. The ViT model was selected as an alternative to convolution-based deep learning models given its success on natural image tasks^31^ and its increasing adoption in medical imaging-related tasks^32^. Both model architectures are available in most of the deep learning frameworks with and without supervised pre-trained weights on ImageNet, which is beneficial when training data is scarce. However, supervised pre-training does not always benefit the downstream task, with the pre-training dataset and objective having an impact on the final performance of the fine-tuned model^33^. For this reason, self-supervised contrastive pre-training^25^ was employed to bridge the gap between the pre-training and downstream dataset and objective. Overall, classification results did not benefit from the contrastive pre-training, with the best-performing model being fine-tuned from ImageNet weights. One reason the anticipated benefits of contrastive learning were not observed could be that models were trained to learn shared information from augmented views of the same transversal slice, discarding the fact that this information should be shared by all the slices in a subject. Nonetheless, PCA of the learned feature space showed a better distinction of the different tumor types when using contrastive pre-training, especially for ResNet50 models.

### MR sequences

Among the MR sequences, ADC achieved the highest overall classification performance and class-wise F1-scores for both ASTR and MED tumor types, whereas models trained on T2w data achieved the highest F1-score for the EP class (see Table S1 in the *Supplementary material*). This can be attributed to the small number of EPs having ADC data (n=9) compared to those having T2w data (n=30). The results on ADC data were in agreement with those using deep-learning based-methods^12^, as well as intensity analysis^34^, and consistent with the information neuro-radiologists use when assessing tumor cellularity and possible tumor grade, during the primary diagnosis work-up.

### Age information

A joint fusion approach was used to combine the image and age information, allowing the image and age encoder to be jointly trained. Results show that the addition of age information did not improve classification performance across the different MR sequences, model architectures, and pre-training strategies. Preliminary investigations also explored the number of encoding layers in the age encoder, with no variation in outcome. This can be attributed to the overlapping age distribution of the different classes as well as to the choice of data fusion approach. By contrast, in a similar experimental setting the combination of image and age information improved model classification performance^12^. Thus, this leaves open the question of whether the benefits of combining age and image information for pediatric brain tumor classification are restricted to specific subject populations or if a more general method for image and age fusion needs to be explored which can be broadly and successfully applied.

### Model explainability

To qualitatively assess the regions used for predictions class-activation mapping was implemented to highlight the regions in the image used by the model for prediction. Given the depth and complexity of both ResNet50 and ViT models, activation maps do not have sufficient spatial resolution to target the tumor region only and provide a visualization of tumor regions and/or image features that are relevant for the classification. Nevertheless, the models’ activation maps showed that the information used for classification fell within the brain region. Interestingly, the effect of the pre-training strategy and the input configuration seen in the feature space visualizations does not reflect on the class activation maps, suggesting that the models use the same brain regions for classification but rely on a somewhat different set of features.

### Comparison with related work

The findings align with the few previously reported studies on deep learning-based pediatric brain tumor type classification, with Quon *et al*.,^11^ reporting a 0.92 accuracy (F1-score of 0.80 on T2w data classifying 4 tumor types and controls) and Artzi *et al*.,^12^ of 0.87 (F1-score 0.82 on diffusion Trace data classifying 3 tumor types and controls). This study’s class-wise results for ASTR and MB are similar to those previously reported, with EP having the lowest score in all studies. It should be noted that the comparison of the performance metrics among the studies can only be considered in very general terms due to the differences in tumor types included in the analysis and the evaluation protocol. Additionally, previous studies did not provide statistical analysis to assess the impact of specific MR-sequence and/or age fusion on models’ performance.

### Limitations

This study has some limitations, particularly concerning two major aspects: (1) the amount and quality of data and (2) the use of 2D models. The dataset, although one of the largest accessible, has a relatively small size and the distribution of tumor types is unbalanced. Not all tumor types available in the dataset could be included in the analysis, limiting the applicability of the trained models in a real-world clinical scenario. The image quality varied greatly between each scan and subject. This large variability in data quality is advantageous for the development of a robust classification method but also adds complexity in determining which factors to adjust to optimize model performance. Moreover, the information regarding the site where the scans were acquired was missing. If available, it could have been used to stratify the subjects in training and testing sets based on MR-site. Additionally, we have only explored the use of single MR sequences separately, while multiple MR sequences (*e*.*g*., T2w and ADC data) should be considered in the future. Finally, the choice of 2D deep-learning models instead of 3D ones resulted in the spatial relationship between the slice and the tridimensionality of the tumor structure being lost. If more data were available, using the MR volumes as model input could address this limitation. Alternatively, while still using 2D slides, aggregation methods that incorporate the slice position for computing volume-wise predictions could be developed. Considering the limitations, the results for this study are preliminary and highlight the need for further research to develop methods that can match the diagnostic performance of radiologists that, for the investigated brain tumors, is 91.7% (range [85.1, 96.7]%) when using T1-w with and without contrast, T2-w, FLAIR and ADC^7^.

## Conclusions

In this proof-of-concept study, the classification of pediatric brain tumors based on MR-images was achieved using deep learning methods. The vision transformer model pre-trained on ImageNet and fine-tuned on ADC data obtained the highest classification performance, with models trained on T2w data also achieving reasonable performance. Image and age fusion did improve classification performance, but not significantly. In future studies, the combination of multiple MR sequences along with more detailed clinical information and further refinements of the network architectures, pre-training and data fusion are warranted o aid radiologists in the clinical assessment of these tumors.

## Data Availability

All data used in this study is available upon reasonable request to The Children's Brain Tumor Tissue Consortium (CBTTC) / The children's brain tumor network (CBTN).
https://cbtn.org/

## Acknowledgments

The research was made possible in part due to The Children’s Brain Tumor Tissue Consortium (CBTTC) / The children’s brain tumor network (CBTN). The study was financed by Swedish Childhood Cancer Foundation (MT2021-0011, MT2022-0013), Joanna Cocozza’s Foundation (2022-2024), Linköping University’s Cancer Strength Area (2022) and ALF Grants, Region Östergötland (974566).

## 1 Code availability

Code linked to this manuscript is available at https://github.com/IulianEmilTampu/PediatricBrainTumorClassification.

## Supplementary material

### Material and Methods

#### ADC computation

The DW-MR data was processed using MRtrix3 software^1^ to obtain a diffusion tensor from which the ADC map was calculated. Briefly, Gibbs-ringing artefact removal, denoising and motion correction were performed before the diffusion tensor fitting. The ADC map was then computed.

#### Intensity normalization, volume re-sampling and slice selection

Data harmonization was performed since the CBTN dataset was obtained from a variety of centres and MRI scanners^2^. The brain from each MRI volume was extracted using a deep learning-based brain extraction tool^3^, followed by a per-sequence voxel intensity clipping confining the values in the [0.2^th^, 99.8^th^] percentile range of the brain region only. Min-max intensity normalization was also performed bringing the voxel intensity values in the [0, 1] range. Lastly, each volume was isotropically interpolated to 1 mm isotropic resolution and reshaped to have 224 × 224 pixels in the transverse plane using an order five spline interpolation function (nibabel.processing.conform).

#### Data splitting

Scans were split subject-wise, into training and validation (80%) and testing (20%), ensuring that subjects with multiple scans did not end up in the same set. When using pre-trained weights from the *in-domain* data (CBTN), test subjects were not used for neither for pre-training nor during fine-tuning, resulting in each of the ten repetitions having a different set of pre-training weights.

#### Pre-training

Transfer learning has been widely investigated and used to address the data scarcity problem in medical image analysis^4^. Thus, in this study, the image feature extractors underwent fine-tuning using pre-trained weights obtained from three distinct pre-training strategies: (1) supervised pre-training on *out-of-domain* data (ImageNet1K), (2) self-supervised pre-training on *close-to-domain* data (BraTS) and (3) self-supervised pre-training on *in-domain* (CBTN) data. Specifically, supervised pre-training utilized ImageNet1K dataset which is a collection of 1.2 million images divided in 1000 classes and is broadly used in literature for model pre-training and with pre-trained model weights available for download from most of the deep learning frameworks (ResNet50 and ViT ImageNet1K model weights from Pytorch were used in this study). For the self-supervised pre-training, the SimCLR^5^ framework was employed, implementing contrastive learning which enables models to learn visual representation from the data without the need of labels. Two distinct datasets were used for self-supervised pre-training: a *close-to-domain* dataset comprising of transversal slices of adult brain tumor obtained from BraTS2020^6–8^, and an *in-domain* dataset composed of transversal slices from the CBTN dataset including all brain images showing tumor. Models were pre-trained on T2-w (TCGA n=22811, CBTN n=5803), T1w-Gd (TCGA n=22811, CBTN n=2584) or ADC (CBTN n=1383) images separately, to match the MR-sequences available for the CBTN dataset. Since BraTS2020 dataset does not provide ADC data, pre-trained weights on T2-w images were used when fine-tuning on the ADC target dataset.

#### Image and age data fusion

Several data fusion approaches have been proposed^9–11^, such as early fusion, joint fusion and late fusion. In this work, joint fusion (feature fusion) was used to combine image and age information for the prediction of tumor type. The advantage of joint fusion is that a single model is trained on both image and age information, with the age not blended immediately with the risk of not being properly used^11^. The age was encoded using a one dense layer bringing the age information into a three-valued vector. The encoded age vector was then concatenated to the encoded image vector before being fed to the classifier part of the model.

#### Implementation and training

Models were implemented in Python (3.9.17) using the PyTorch^12^ framework (2.0.1+cu117), and were trained on a computer with 20-core CPU and 4 NVIDIA Tesla V100 GPUs (32GB memory each). SimCLR pre-training on both the *close-to-domain* and *in-domain* data run for 500 epochs, minimizing the contrastive loss between positive pairs of augmented images (see^5^ for details) using AdamW optimizer^13^ with CosineAnnealingLR learning rate scheduler^14^ (initial learning rate=1.0e-05). During fine-tuning on the target CBTN dataset, only the last 2 convolutional blocks of ResNet50 (22.1M parameters) and the last 6 attention layers for ViT (43.3M parameters) were trained to minimize the weighted categorical cross-entropy loss with balanced weights computed on the training set (sklearn.utils.class_weight function). Adam optimizer^15^ (*β*_1_ = 0.9, *β*_2_ = 0.999) with exponential learning rate decay (starting learning rate=1e-5, *γ* = 0.99) was also used. Fine-tuning run for 200 epochs with the training stopping if the validation loss did not decrease over 20 epochs.

Data augmentation was applied^16–19^ during both pre-training and fine-tuning, using both geometric transformations (random rotations between ±45^°^, random horizontal and vertical flipping, and 10% random width and height shift) and random color jitter (brightness up to 50%). In addition, the TrivialWideAugment automatic data augmentation method^20^ was also used, given negligible computational overhead and performance improvement see in natural image classification tasks (num_magnitude_bins=15). Code linked to this manuscript is available at (removed for anonymity).

**Figure S1.**
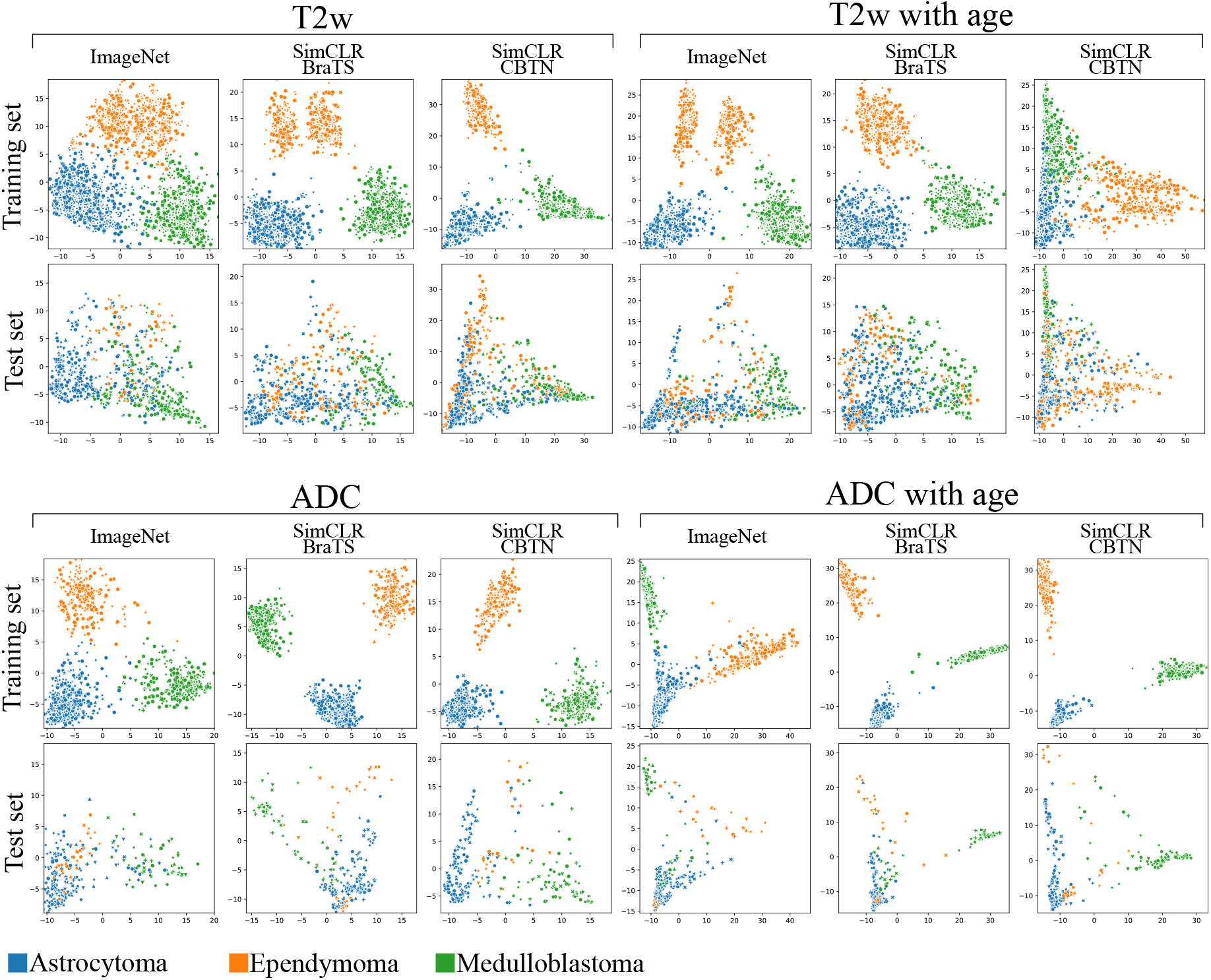
Principal component analysis (PCA) of image features extracted by ResNet50 models fine-tuned on T2w or ADC data, with and without age information, using ImageNet or SimCLR pre-trained weights. The first and second principal components are presented, for both training and testing sets. Classes are color-coded.

## Results

**Table S1.** Subjects-wise classification performance across all the available MR sequences (alone or with age fusion), for the implemented deep learning models and pre-training strategy. Classe-wise metrics are also provided. std: standard deviation, MCC: Matthew’s correlation coefficient, AUC: area under the ROC curve (marco-average). Table continues on multiple pages.

| MR sequence | Model version | Use age | Pre-training strategy | Pre-training dataset | MCC [mean±std] | Accuracy [mean±std] | AUC [mean±std] | Tumor type | Precision [mean±std] | Recall [mean±std] | F1-score [mean±std] |
| --- | --- | --- | --- | --- | --- | --- | --- | --- | --- | --- | --- |
| T1 | ResNet50 | False | / | Imagenet | 0.3836 ± 0.0961 | 0.5946 ± 0.0751 | 0.7632 ± 0.0571 | ASTR | 0.8494 ± 0.0897 | 0.5325 ± 0.1906 | 0.5614 ± 0.1544 |
|  |  |  |  |  |  |  |  | EP | 0.8582 ± 0.0966 | 0.3037 ± 0.1985 | 0.3319 ± 0.2153 |
|  |  |  |  |  |  |  |  | MED | 0.6842 ± 0.1290 | 0.7905 ± 0.1628 | 0.6850 ± 0.1106 |
|  |  |  | SimCLR | TCGA | 0.3733 ± 0.1286 | 0.6051 ± 0.0856 | 0.7404 ± 0.1089 | ASTR | 0.8008 ± 0.0939 | 0.5372 ± 0.1582 | 0.5496 ± 0.1158 |
|  |  |  |  |  |  |  |  | EP | 0.8953 ± 0.0748 | 0.3030 ± 0.1831 | 0.3523 ± 0.1993 |
|  |  |  |  |  |  |  |  | MED | 0.6794 ± 0.0999 | 0.8008 ± 0.1381 | 0.7032 ± 0.1164 |
|  |  |  |  | CBTN | 0.3875 ± 0.1316 | 0.6095 ± 0.0853 | 0.7581 ± 0.0884 | ASTR | 0.7650 ± 0.1027 | 0.5851 ± 0.1224 | 0.5657 ± 0.1201 |
|  |  |  |  |  |  |  |  | EP | 0.9043 ± 0.0629 | 0.3127 ± 0.2007 | 0.3533 ± 0.2010 |
|  |  |  |  |  |  |  |  | MED | 0.7310 ± 0.0986 | 0.7880 ± 0.1217 | 0.7137 ± 0.1047 |
|  |  | True | / | Imagenet | 0.3513 ± 0.1558 | 0.5735 ± 0.1078 | 0.7436 ± 0.0869 | ASTR | 0.8464 ± 0.0960 | 0.4818 ± 0.1945 | 0.5220 ± 0.1824 |
|  |  |  |  |  |  |  |  | EP | 0.8332 ± 0.1250 | 0.3060 ± 0.2073 | 0.3187 ± 0.2079 |
|  |  |  |  |  |  |  |  | MED | 0.6757 ± 0.1272 | 0.7812 ± 0.1983 | 0.6774 ± 0.1356 |
|  |  |  | SimCLR | TCGA | 0.3957 ± 0.1184 | 0.6160 ± 0.0828 | 0.7546 ± 0.0993 | ASTR | 0.8190 ± 0.1070 | 0.5458 ± 0.1642 | 0.5618 ± 0.1161 |
|  |  |  |  |  |  |  |  | EP | 0.9025 ± 0.0757 | 0.3027 ± 0.1722 | 0.3550 ± 0.1725 |
|  |  |  |  |  |  |  |  | MED | 0.6756 ± 0.1214 | 0.8232 ± 0.1282 | 0.7131 ± 0.1164 |
|  |  |  |  | CBTN | 0.4124 ± 0.1123 | 0.6235 ± 0.0799 | 0.7741 ± 0.0759 | ASTR | 0.8041 ± 0.0931 | 0.5813 ± 0.1444 | 0.5788 ± 0.1264 |
|  |  |  |  |  |  |  |  | EP | 0.8987 ± 0.0792 | 0.3003 ± 0.2129 | 0.3391 ± 0.2197 |
|  |  |  |  |  |  |  |  | MED | 0.7244 ± 0.1142 | 0.8159 ± 0.1047 | 0.7269 ± 0.1077 |
|  | ViT_b_16 | False | / | Imagenet | 0.3307 ± 0.1208 | 0.5838 ± 0.0787 | 0.6925 ± 0.1006 | ASTR | 0.8023 ± 0.0913 | 0.5337 ± 0.1782 | 0.5467 ± 0.1713 |
|  |  |  |  |  |  |  |  | EP | 0.9337 ± 0.0650 | 0.2190 ± 0.1877 | 0.2736 ± 0.2151 |
|  |  |  |  |  |  |  |  | MED | 0.5849 ± 0.1099 | 0.7897 ± 0.1675 | 0.6545 ± 0.1101 |
|  |  |  | SimCLR | TCGA | 0.4540 ± 0.1557 | 0.6585 ± 0.0993 | 0.7696 ± 0.1028 | ASTR | 0.8071 ± 0.0892 | 0.6511 ± 0.1882 | 0.6346 ± 0.1235 |
|  |  |  |  |  |  |  |  | EP | 0.9321 ± 0.0648 | 0.3063 ± 0.2244 | 0.3754 ± 0.2565 |
|  |  |  |  |  |  |  |  | MED | 0.7079 ± 0.1699 | 0.8313 ± 0.1252 | 0.7416 ± 0.0942 |
|  |  |  |  | CBTN | 0.3763 ± 0.0981 | 0.6168 ± 0.0673 | 0.7376 ± 0.1036 | ASTR | 0.7690 ± 0.0898 | 0.6033 ± 0.1543 | 0.5831 ± 0.1215 |
|  |  |  |  |  |  |  |  | EP | 0.9415 ± 0.0547 | 0.1953 ± 0.1891 | 0.2576 ± 0.2335 |
|  |  |  |  |  |  |  |  | MED | 0.6597 ± 0.1355 | 0.8099 ± 0.1255 | 0.7094 ± 0.1101 |
|  |  | True | / | Imagenet | 0.3276 ± 0.1154 | 0.5793 ± 0.0824 | 0.6925 ± 0.0879 | ASTR | 0.8012 ± 0.1078 | 0.5377 ± 0.1605 | 0.5512 ± 0.1482 |
|  |  |  |  |  |  |  |  | EP | 0.9269 ± 0.0775 | 0.2077 ± 0.1922 | 0.2670 ± 0.2254 |
|  |  |  |  |  |  |  |  | MED | 0.5907 ± 0.0906 | 0.7808 ± 0.1840 | 0.6491 ± 0.1218 |
|  |  |  | SimCLR | TCGA | 0.4383 ± 0.1250 | 0.6466 ± 0.0840 | 0.7645 ± 0.0997 | ASTR | 0.7970 ± 0.1003 | 0.6295 ± 0.1685 | 0.6168 ± 0.1019 |
|  |  |  |  |  |  |  |  | EP | 0.9260 ± 0.0670 | 0.3067 ± 0.2279 | 0.3697 ± 0.2604 |
|  |  |  |  |  |  |  |  | MED | 0.7052 ± 0.1712 | 0.8210 ± 0.1347 | 0.7342 ± 0.0971 |
|  |  |  |  | CBTN | 0.3522 ± 0.1077 | 0.6008 ± 0.0720 | 0.7247 ± 0.0992 | ASTR | 0.7528 ± 0.1052 | 0.6235 ± 0.1775 | 0.5877 ± 0.1319 |
|  |  |  |  |  |  |  |  | EP | 0.9424 ± 0.0493 | 0.1590 ± 0.1742 | 0.2070 ± 0.2209 |
|  |  |  |  |  |  |  |  | MED | 0.6497 ± 0.1495 | 0.7865 ± 0.1353 | 0.6857 ± 0.0915 |
| T2 | ResNet50 | False | / | ImageNet | 0.5205 ± 0.1242 | 0.6835 ± 0.0830 | 0.8534 ± 0.0470 | ASTR | 0.8716 ± 0.0622 | 0.7260 ± 0.1307 | 0.7703 ± 0.0905 |
|  |  |  |  |  |  |  |  | EP | 0.8668 ± 0.0656 | 0.4374 ± 0.2270 | 0.4166 ± 0.2088 |
|  |  |  |  |  |  |  |  | MED | 0.7994 ± 0.1080 | 0.8053 ± 0.1283 | 0.7083 ± 0.0636 |
|  |  |  | SimCLR | TCGA | 0.5475 ± 0.1057 | 0.7016 ± 0.0734 | 0.8531 ± 0.0492 | ASTR | 0.8692 ± 0.0669 | 0.7036 ± 0.1245 | 0.7574 ± 0.1022 |
|  |  |  |  |  |  |  |  | EP | 0.8930 ± 0.0658 | 0.5141 ± 0.2043 | 0.5088 ± 0.1624 |
|  |  |  |  |  |  |  |  | MED | 0.7942 ± 0.0888 | 0.8388 ± 0.1089 | 0.7237 ± 0.0734 |
|  |  |  |  | CBTN | 0.5453 ± 0.1112 | 0.6997 ± 0.0765 | 0.8546 ± 0.0491 | ASTR | 0.8673 ± 0.0765 | 0.7065 ± 0.1436 | 0.7564 ± 0.1179 |
|  |  |  |  |  |  |  |  | EP | 0.8827 ± 0.0570 | 0.5034 ± 0.1895 | 0.4910 ± 0.1546 |
|  |  |  |  |  |  |  |  | MED | 0.8056 ± 0.0972 | 0.8419 ± 0.1113 | 0.7334 ± 0.0673 |
|  |  | True | / | ImageNet | 0.5281 ± 0.1260 | 0.6862 ± 0.0902 | 0.8464 ± 0.0547 | ASTR | 0.8857 ± 0.0690 | 0.7103 ± 0.1583 | 0.7623 ± 0.1432 |
|  |  |  |  |  |  |  |  | EP | 0.8640 ± 0.0761 | 0.4743 ± 0.2344 | 0.4395 ± 0.1938 |
|  |  |  |  |  |  |  |  | MED | 0.7934 ± 0.1052 | 0.8087 ± 0.1258 | 0.7075 ± 0.0750 |
|  |  |  | SimCLR | TCGA | 0.5591 ± 0.0984 | 0.7105 ± 0.0673 | 0.8474 ± 0.0565 | ASTR | 0.8732 ± 0.0648 | 0.7188 ± 0.1203 | 0.7693 ± 0.0990 |
|  |  |  |  |  |  |  |  | EP | 0.9042 ± 0.0678 | 0.5084 ± 0.1717 | 0.5200 ± 0.1549 |
|  |  |  |  |  |  |  |  | MED | 0.7916 ± 0.0938 | 0.8390 ± 0.0996 | 0.7245 ± 0.0788 |
|  |  |  |  | CBTN | 0.5410 ± 0.1084 | 0.6946 ± 0.0801 | 0.8456 ± 0.0539 | ASTR | 0.8731 ± 0.0634 | 0.6799 ± 0.1639 | 0.7345 ± 0.1492 |
|  |  |  |  |  |  |  |  | EP | 0.8816 ± 0.0678 | 0.5246 ± 0.1916 | 0.5035 ± 0.1519 |
|  |  |  |  |  |  |  |  | MED | 0.7946 ± 0.1001 | 0.8466 ± 0.1040 | 0.7307 ± 0.0772 |
|  | ViT_b_16 | False | / | ImageNet | 0.5458 ± 0.1276 | 0.7097 ± 0.0836 | 0.8387 ± 0.0533 | ASTR | 0.8415 ± 0.0826 | 0.7829 ± 0.1245 | 0.7964 ± 0.1009 |
|  |  |  |  |  |  |  |  | EP | 0.9056 ± 0.0557 | 0.4273 ± 0.2024 | 0.4543 ± 0.1812 |
|  |  |  |  |  |  |  |  | MED | 0.8068 ± 0.0726 | 0.8030 ± 0.1223 | 0.7149 ± 0.0577 |
|  |  |  | SimCLR | TCGA | 0.5687 ± 0.0987 | 0.7232 ± 0.0623 | 0.8376 ± 0.0548 | ASTR | 0.8375 ± 0.0832 | 0.7972 ± 0.1012 | 0.8034 ± 0.0670 |
|  |  |  |  |  |  |  |  | EP | 0.9167 ± 0.0514 | 0.4733 ± 0.2070 | 0.4905 ± 0.1758 |
|  |  |  |  |  |  |  |  | MED | 0.8229 ± 0.0840 | 0.7917 ± 0.1039 | 0.7176 ± 0.0781 |
|  |  |  |  | CBTN | 0.5504 ± 0.1081 | 0.7098 ± 0.0732 | 0.8270 ± 0.0561 | ASTR | 0.8174 ± 0.0945 | 0.7737 ± 0.1204 | 0.7788 ± 0.0888 |
|  |  |  |  |  |  |  |  | EP | 0.9306 ± 0.0516 | 0.4546 ± 0.1953 | 0.4884 ± 0.1682 |
|  |  |  |  |  |  |  |  | MED | 0.8033 ± 0.0934 | 0.8051 ± 0.1365 | 0.7091 ± 0.0742 |
|  |  | True | / | ImageNet | 0.5612 ± 0.1132 | 0.7174 ± 0.0774 | 0.8507 ± 0.0482 | ASTR | 0.8466 ± 0.0776 | 0.7844 ± 0.1240 | 0.7979 ± 0.0916 |
|  |  |  |  |  |  |  |  | EP | 0.9141 ± 0.0507 | 0.4295 ± 0.2100 | 0.4546 ± 0.1836 |
|  |  |  |  |  |  |  |  | MED | 0.8077 ± 0.0871 | 0.8280 ± 0.1080 | 0.7300 ± 0.0632 |
|  |  |  | SimCLR | TCGA | 0.5798 ± 0.1141 | 0.7307 ± 0.0752 | 0.8459 ± 0.0556 | ASTR | 0.8283 ± 0.0871 | 0.7944 ± 0.0952 | 0.7980 ± 0.0756 |
|  |  |  |  |  |  |  |  | EP | 0.9231 ± 0.0464 | 0.4847 ± 0.2123 | 0.5098 ± 0.1833 |
|  |  |  |  |  |  |  |  | MED | 0.8327 ± 0.0857 | 0.8140 ± 0.1239 | 0.7384 ± 0.0803 |
|  |  |  |  | CBTN | 0.5259 ± 0.1747 | 0.6966 ± 0.1163 | 0.8096 ± 0.0756 | ASTR | 0.7934 ± 0.1478 | 0.7594 ± 0.1418 | 0.7599 ± 0.1126 |
|  |  |  |  |  |  |  |  | EP | 0.9294 ± 0.1231 | 0.4488 ± 0.2152 | 0.4955 ± 0.2069 |
|  |  |  |  |  |  |  |  | MED | 0.8020 ± 0.1019 | 0.7859 ± 0.1968 | 0.6836 ± 0.1586 |
| ADC | ResNet50 | False | / | ImageNet | 0.6708 ± 0.1440 | 0.8127 ± 0.0830 | 0.9096 ± 0.0800 | ASTR | 0.7873 ± 0.1416 | 0.8757 ± 0.1121 | 0.8591 ± 0.0671 |
|  |  |  |  |  |  |  |  | EP | 0.9090 ± 0.0778 | 0.4222 ± 0.4211 | 0.3326 ± 0.3175 |
|  |  |  |  |  |  |  |  | MED | 0.9668 ± 0.0590 | 0.8533 ± 0.1542 | 0.8674 ± 0.1055 |
|  |  |  | SimCLR | TCGA | 0.7045 ± 0.1180 | 0.8428 ± 0.0579 | 0.9434 ± 0.0631 | ASTR | 0.7723 ± 0.1363 | 0.9375 ± 0.0631 | 0.8986 ± 0.0469 |
|  |  |  |  |  |  |  |  | EP | 0.9421 ± 0.0608 | 0.2556 ± 0.3890 | 0.2296 ± 0.3339 |
|  |  |  |  |  |  |  |  | MED | 0.9744 ± 0.0505 | 0.8817 ± 0.1313 | 0.8870 ± 0.0889 |
|  |  |  |  | CBTN | 0.7238 ± 0.1261 | 0.8556 ± 0.0557 | 0.9308 ± 0.0860 | ASTR | 0.7341 ± 0.1644 | 0.9606 ± 0.0578 | 0.9044 ± 0.0490 |
|  |  |  |  |  |  |  |  | EP | 0.9522 ± 0.0558 | 0.3333 ± 0.4082 | 0.2926 ± 0.3336 |
|  |  |  |  |  |  |  |  | MED | 0.9937 ± 0.0249 | 0.8400 ± 0.1615 | 0.8941 ± 0.1043 |
|  |  | True | / | ImageNet | 0.5831 ± 0.2263 | 0.7427 ± 0.1627 | 0.8904 ± 0.1450 | ASTR | 0.8000 ± 0.1588 | 0.7599 ± 0.2305 | 0.7766 ± 0.1963 |
|  |  |  |  |  |  |  |  | EP | 0.8657 ± 0.1370 | 0.4000 ± 0.4295 | 0.2755 ± 0.3200 |
|  |  |  |  |  |  |  |  | MED | 0.9208 ± 0.1397 | 0.8450 ± 0.2088 | 0.8272 ± 0.1990 |
|  |  |  | SimCLR | TCGA | 0.7100 ± 0.1357 | 0.8405 ± 0.0767 | 0.9409 ± 0.0623 | ASTR | 0.7931 ± 0.1374 | 0.9233 ± 0.0934 | 0.8936 ± 0.0604 |
|  |  |  |  |  |  |  |  | EP | 0.9342 ± 0.0725 | 0.3444 ± 0.4193 | 0.3037 ± 0.3483 |
|  |  |  |  |  |  |  |  | MED | 0.9705 ± 0.0543 | 0.8783 ± 0.1387 | 0.8860 ± 0.0985 |
|  |  |  |  | CBTN | 0.6935 ± 0.1305 | 0.8341 ± 0.0734 | 0.9304 ± 0.0772 | ASTR | 0.7412 ± 0.1510 | 0.9454 ± 0.0863 | 0.8933 ± 0.0588 |
|  |  |  |  |  |  |  |  | EP | 0.9360 ± 0.0780 | 0.2222 ± 0.3583 | 0.1978 ± 0.2975 |
|  |  |  |  |  |  |  |  | MED | 0.9815 ± 0.0433 | 0.8433 ± 0.1560 | 0.8844 ± 0.0969 |
|  | ViT_b_16 | False | / | ImageNet | 0.7673 ± 0.1321 | 0.8721 ± 0.0737 | 0.9310 ± 0.0770 | ASTR | 0.7863 ± 0.1540 | 0.9608 ± 0.0642 | 0.9089 ± 0.0569 |
|  |  |  |  |  |  |  |  | EP | 0.9637 ± 0.0486 | 0.4444 ± 0.4246 | 0.4222 ± 0.3843 |
|  |  |  |  |  |  |  |  | MED | 0.9867 ± 0.0419 | 0.8617 ± 0.1487 | 0.8950 ± 0.0999 |
|  |  |  | SimCLR | TCGA | 0.6827 ± 0.1185 | 0.8268 ± 0.0670 | 0.9442 ± 0.0655 | ASTR | 0.7717 ± 0.1411 | 0.9246 ± 0.0973 | 0.8840 ± 0.0563 |
|  |  |  |  |  |  |  |  | EP | 0.9211 ± 0.0760 | 0.2000 ± 0.3399 | 0.1622 ± 0.2628 |
|  |  |  |  |  |  |  |  | MED | 0.9781 ± 0.0492 | 0.8683 ± 0.1435 | 0.8892 ± 0.0948 |
|  |  |  |  | CBTN | 0.6647 ± 0.1629 | 0.8242 ± 0.0732 | 0.9062 ± 0.0918 | ASTR | 0.7038 ± 0.1986 | 0.9404 ± 0.0964 | 0.8748 ± 0.0633 |
|  |  |  |  |  |  |  |  | EP | 0.9502 ± 0.0689 | 0.0889 ± 0.2183 | 0.0956 ± 0.2353 |
|  |  |  |  |  |  |  |  | MED | 0.9795 ± 0.0457 | 0.8250 ± 0.1635 | 0.8617 ± 0.1141 |
|  |  | True | / | ImageNet | 0.7730 ± 0.1439 | 0.8738 ± 0.0839 | 0.9399 ± 0.0732 | ASTR | 0.7977 ± 0.1527 | 0.9659 ± 0.0630 | 0.9165 ± 0.0600 |
|  |  |  |  |  |  |  |  | EP | 0.9562 ± 0.0673 | 0.4444 ± 0.4246 | 0.4481 ± 0.4080 |
|  |  |  |  |  |  |  |  | MED | 0.9941 ± 0.0233 | 0.8517 ± 0.1557 | 0.9016 ± 0.0955 |
|  |  |  | SimCLR | TCGA | 0.6807 ± 0.1356 | 0.8287 ± 0.0758 | 0.9280 ± 0.0732 | ASTR | 0.7258 ± 0.1337 | 0.9255 ± 0.0997 | 0.8694 ± 0.0656 |
|  |  |  |  |  |  |  |  | EP | 0.9397 ± 0.0628 | 0.1889 ± 0.3213 | 0.1793 ± 0.2916 |
|  |  |  |  |  |  |  |  | MED | 0.9885 ± 0.0360 | 0.8600 ± 0.1541 | 0.8941 ± 0.1021 |
|  |  |  |  | CBTN | 0.6906 ± 0.1394 | 0.8368 ± 0.0696 | 0.9225 ± 0.0894 | ASTR | 0.7272 ± 0.2114 | 0.9495 ± 0.0811 | 0.8908 ± 0.0613 |
|  |  |  |  |  |  |  |  | EP | 0.9438 ± 0.0568 | 0.1889 ± 0.3542 | 0.1519 ± 0.2763 |
|  |  |  |  |  |  |  |  | MED | 0.9873 ± 0.0344 | 0.8267 ± 0.1597 | 0.8761 ± 0.1063 |

**Figure S2.**
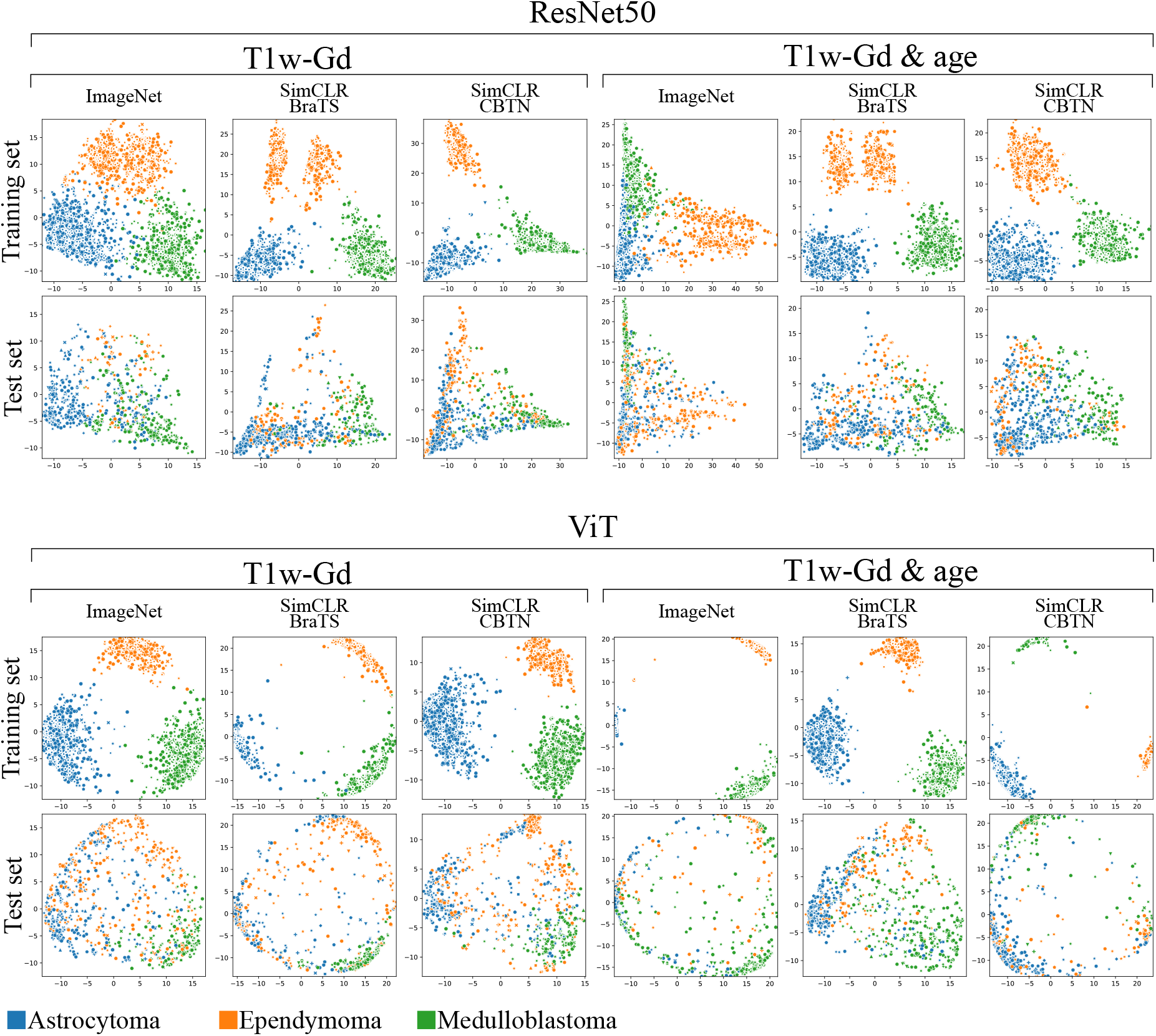
Principal component analysis (PCA) of image features extracted by ResNet50 and ViT models fine-tuned on T1W-Gd data, with and without age information, using ImageNet or SimCLR pre-trained weights. The first and second principal components are presented, for both training and testing sets. Classes are color-coded.

**Figure S3.**
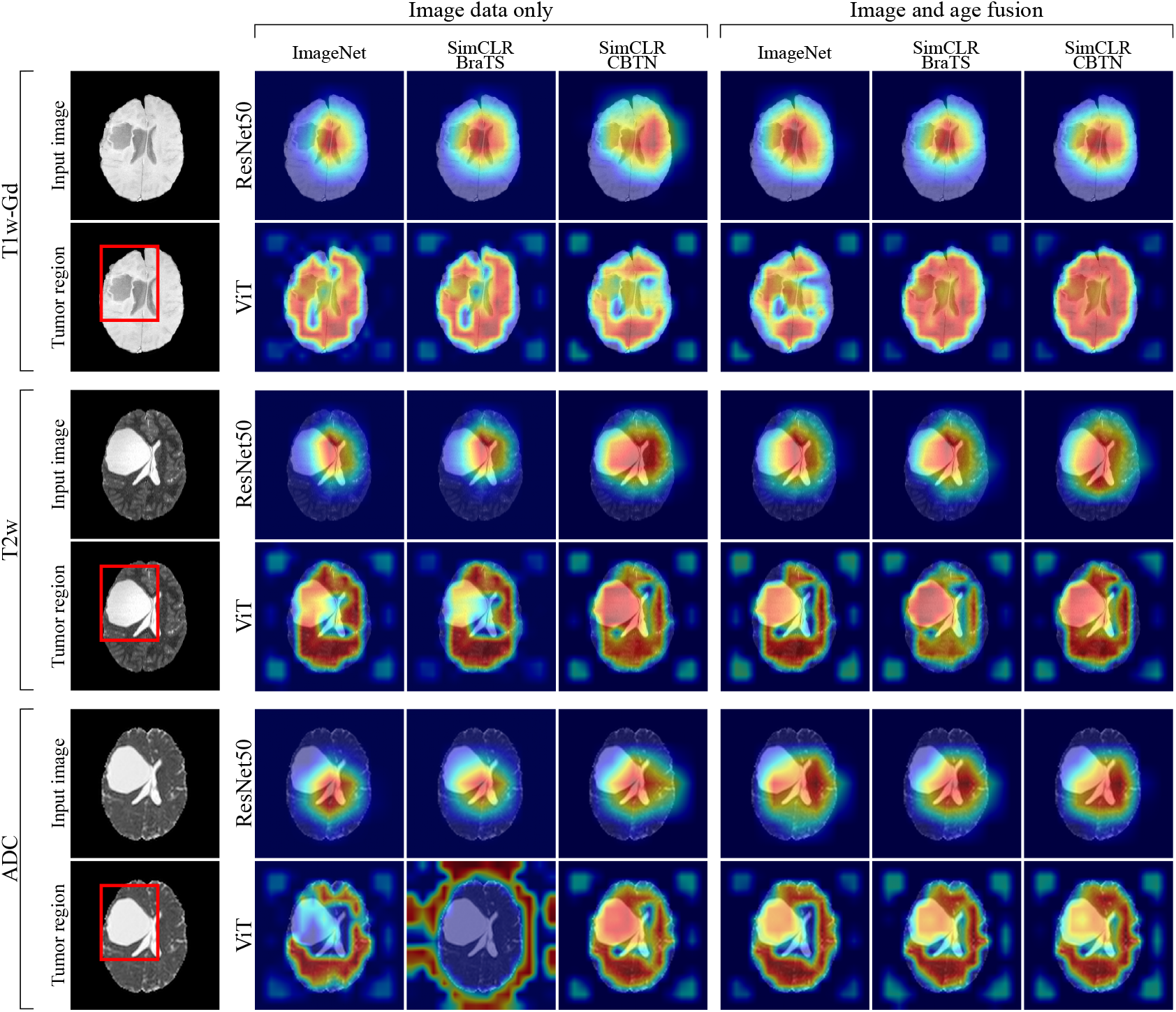
Grad-CAMs for the models trained on different MR-sequences, with and without age fusion, and for the three pre-training strategies investigated. Grad-CAMs are computed with respect to the ground truth. The red square in the *tumor region* panel delineates the tumor. In the Grad-CAMs images, red color identify the parts of the input image used mostly contributing to the classification.

## References

1. Ferlay, J., Ervik, M., Lam, F. et al. Global cancer observatory: cancer today. https://gco.iarc.fr/today/home (2022). Accessed: 2023.

2. Sharma, R. A systematic examination of burden of childhood cancers in 183 countries: estimates from GLOBOCAN 2018. Eur. J. Cancer Care 30, e13438 (2021).

3. Ostrom, Q. T., Patil, N., Cioffi, G. et al. CBTRUS statistical report: primary brain and other central nervous system tumors diagnosed in the United States in 2013–2017. Neuro-oncology 22, iv1–iv96 (2020).

4. Corti, C., Urgesi, C., Massimino, M. et al. Effects of supratentorial and infratentorial tumor location on cognitive functioning of children with brain tumor. Child’s Nerv. Syst. 36, 513–524 (2020).

5. Pollack, I. F. Brain tumors in children. New Engl. J. Medicine 331, 1500–1507 (1994).

6. Louis, D. N., Perry, A., Wesseling, P. et al. The 2021 WHO classification of tumors of the central nervous system: a summary. Neuro-oncology 23, 1231–1251 (2021).

7. Dixon, L., Jandu, G. K., Sidpra, J. & Mankad, K. Diagnostic accuracy of qualitative MRI in 550 paediatric brain tumours: evaluating current practice in the computational era. Quant. Imaging Medicine Surg. 12, 131 (2022).

8. Ali, S., Li, J., Pei, Y. et al. A comprehensive survey on brain tumor diagnosis using deep learning and emerging hybrid techniques with multi-modal MR image. Arch. Comput. Methods Eng. 29, 4871–4896 (2022).

9. Amin, J., Sharif, M., Haldorai, A. et al. Brain tumor detection and classification using machine learning: a comprehensive survey. Complex & Intell. Syst. 1–23 (2021).

10. Tandel, G. S., Biswas, M., Kakde, O. G. et al. A review on a deep learning perspective in brain cancer classification. Cancers 11, 111 (2019).

11. Quon, J., Bala, W., Chen, L. et al. Deep learning for pediatric posterior fossa tumor detection and classification: a multi-institutional study. Am. J. Neuroradiol. 41, 1718–1725 (2020).

12. Artzi, M., Redmard, E., Tzemach, O. et al. Classification of pediatric posterior fossa tumors using convolutional neural network and tabular data. IEEE Access 9, 91966–91973 (2021).

13. Shaari, H., Kevrić, J., Jukić, S. et al. Deep learning-based studies on pediatric brain tumors imaging: narrative review of techniques and challenges. Brain Sci. 11, 716 (2021).

14. Huang, J., Shlobin, N. A., Lam, S. K. & DeCuypere, M. Artificial intelligence applications in pediatric brain tumor imaging: A systematic review. World neurosurgery 157, 99–105 (2022).

15. The Children’s Brain Tumor Network. https://cbtn.org/. Accessed: 2021.

16. Lilly, J. V., Rokita, J. L., Mason, J. L. et al. The children’s brain tumor network (CBTN)-Accelerating research in pediatric central nervous system tumors through collaboration and open science. Neoplasia 35, 100846 (2023).

17. Menze, B. H., Jakab, A., Bauer, S. et al. The multimodal brain tumor image segmentation benchmark (BRATS). IEEE transactions on medical imaging 34, 1993–2024 (2014).

18. Bakas, S., Akbari, H., Sotiras, A. et al. Advancing The Cancer Genome Atlas glioma MRI collections with expert segmentation labels and radiomic features. scientific data 4 (1), 170117 (2017).

19. Bakas, S., Reyes, M., Jakab, A. et al. Identifying the best machine learning algorithms for brain tumor segmentation, progression assessment, and overall survival prediction in the BRATS challenge. preprint 1811.02629 (2018).

20. Tournier, J.-D., Smith, R., Raffelt, D. et al. MRtrix3: A fast, flexible and open software framework for medical image processing and visualisation. Neuroimage 202, 116137 (2019).

21. He, K., Zhang, X., Ren, S. & Sun, J. Deep residual learning for image recognition. In Proceedings of the IEEE conference on computer vision and pattern recognition, 770–778 (2016).

22. Dosovitskiy, A., Beyer, L., Kolesnikov, A. et al. An image is worth 16×16 words: Transformers for image recognition at scale. arXiv preprint 2010.11929 (2020).

23. Bhojanapalli, S., Chakrabarti, A., Glasner, D. et al. Understanding robustness of transformers for image classification. In Proceedings of the IEEE/CVF international conference on computer vision, 10231–10241 (2021).

24. Deng, J., Dong, W., Socher, R. et al. Imagenet: A large-scale hierarchical image database. In 2009 IEEE conference on computer vision and pattern recognition, 248–255 (Ieee, 2009).

25. Chen, T., Kornblith, S., Norouzi, M. & Hinton, G. A simple framework for contrastive learning of visual representations. In International conference on machine learning, 1597–1607 (PMLR, 2020).

26. Chicco, D. & Jurman, G. The advantages of the Matthews correlation coefficient (MCC) over F1 score and accuracy in binary classification evaluation. BMC genomics 21, 1–13 (2020).

27. Zhou, B., Khosla, A., Lapedriza, A. et al. Learning deep features for discriminative localization. In Proceedings of the IEEE conference on computer vision and pattern recognition, 2921–2929 (2016).

28. Selvaraju, R. R., Cogswell, M., Das, A. et al. Grad-CAM: Visual explanations from deep networks via gradient-based localization. In Proceedings of the IEEE international conference on computer vision, 618–626 (2017).

29. Chelghoum, R., Ikhlef, A., Hameurlaine, A. & Jacquir, S. Transfer learning using convolutional neural network architectures for brain tumor classification from MRI images. In Artificial Intelligence Applications and Innovations: 16th IFIP WG 12.5 International Conference, AIAI 2020, Neos Marmaras, Greece, June 5–7, 2020, Proceedings, Part I 16, 189–200 (Springer, 2020).

30. Rehman, A., Naz, S., Razzak, M. I. et al. A deep learning-based framework for automatic brain tumors classification using transfer learning. Circuits, Syst. Signal Process. 39, 757–775 (2020).

31. Khan, S., Naseer, M., Hayat, M. et al. Transformers in vision: A survey. ACM computing surveys (CSUR) 54, 1–41 (2022).

32. Shamshad, F., Khan, S., Zamir, S. W. et al. Transformers in medical imaging: A survey. Med. Image Analysis 102802 (2023).

33. Zoph, B., Ghiasi, G., Lin, T.-Y. et al. Rethinking pre-training and self-training. Adv. neural information processing systems 33, 3833–3845 (2020).

34. Tanyel, T., Nadarajan, C., Duc, N. M. & Keserci, B. Deciphering machine learning decisions to distinguish between posterior fossa tumor types using MRI features: What do the data tell us? Cancers 15, 4015 (2023).

## References

1. Tournier, J.-D., Smith, R., Raffelt, D. et al. MRtrix3: A fast, flexible and open software framework for medical image processing and visualisation. Neuroimage 202, 116137 (2019).

2. Lilly, J. V., Rokita, J. L., Mason, J. L. et al. The children’s brain tumor network (CBTN)-Accelerating research in pediatric central nervous system tumors through collaboration and open science. Neoplasia 35, 100846 (2023).

3. Isensee, F., Schell, M., Pflueger, I. et al. Automated brain extraction of multisequence MRI using artificial neural networks. Hum. brain mapping 40, 4952–4964 (2019).

4. Kim, H. E., Cosa-Linan, A., Santhanam, N. et al. Transfer learning for medical image classification: a literature review. BMC medical imaging 22, 69 (2022).

5. Chen, T., Kornblith, S., Norouzi, M. & Hinton, G. A simple framework for contrastive learning of visual representations. In International conference on machine learning, 1597–1607 (PMLR, 2020).

6. Menze, B. H., Jakab, A., Bauer, S. et al. The multimodal brain tumor image segmentation benchmark (BRATS). IEEE transactions on medical imaging 34, 1993–2024 (2014).

7. Bakas, S., Akbari, H., Sotiras, A. et al. Advancing The Cancer Genome Atlas glioma MRI collections with expert segmentation labels and radiomic features. scientific data 4 (1), 170117 (2017).

8. Bakas, S., Reyes, M., Jakab, A. et al. Identifying the best machine learning algorithms for brain tumor segmentation, progression assessment, and overall survival prediction in the BRATS challenge. preprint 1811.02629 (2018).

9. Acosta, J. N., Falcone, G. J., Rajpurkar, P. & Topol, E.J. Multimodal biomedical AI. Nat. Medicine 28, 1773–1784 (2022).

10. Kline, A., Wang, H., Li, Y. et al. Multimodal machine learning in precision health: A scoping review. npj Digit. Medicine 5, 171 (2022).

11. Huang, S.-C., Pareek, A., Seyyedi, S. et al. Fusion of medical imaging and electronic health records using deep learning: a systematic review and implementation guidelines. NPJ digital medicine 3, 136 (2020).

12. Paszke, A., Gross, S., Massa, F. et al. Pytorch: An imperative style, high-performance deep learning library. Adv. neural information processing systems 32 (2019).

13. Loshchilov, I. & Hutter, F. Decoupled weight decay regularization. arXiv preprint 1711.05101 (2017).

14. Loshchilov, I. & Hutter, F. Sgdr: Stochastic gradient descent with warm restarts. arXiv preprint 1608.03983 (2016).

15. Kingma, D. P. & Ba, J. Adam: A method for stochastic optimization. preprint 1412.6980 (2014).

16. Paul, J. S., Plassard, A. J., Landman, B. A. & Fabbri, D. Deep learning for brain tumor classification. In Medical Imaging 2017: Biomedical Applications in Molecular, Structural, and Functional Imaging, vol. 10137, 253–268 (SPIE, 2017).

17. Chlap, P., Min, H., Vandenberg, N. et al. A review of medical image data augmentation techniques for deep learning applications. J. Med. Imaging Radiat. Oncol. 65, 545–563 (2021).

18. Kumar, R. L., Kakarla, J., Isunuri, B. V. & Singh, M. Multi-class brain tumor classification using residual network and global average pooling. Multimed. Tools Appl. 80, 13429–13438 (2021).

19. Sajjad, M., Khan, S., Muhammad, K. et al. Multi-grade brain tumor classification using deep CNN with extensive data augmentation. J. computational science 30, 174–182 (2019).

20. Müller, S. G. & Hutter, F. Trivialaugment: Tuning-free yet state-of-the-art data augmentation. In Proceedings of the IEEE/CVF international conference on computer vision, 774–782 (2021).

